# Impact of long-lasting insecticidal nets on *Plasmodium falciparum* infection among women attending antenatal clinic at the Bonassama District Hospital, Littoral Region of Cameroon

**DOI:** 10.1101/2024.05.29.24308147

**Authors:** Marcelus U. Ajonina, Irene U. Ajonina-Ekoti, John Ngulefac, Nicholas Ade, Derick N. Awambeng, Carine K. Nfor, Martin Ayim, Tobias O. Apinjoh

## Abstract

Malaria during pregnancy continues to be a significant cause of morbidity and mortality for both infants and mothers, particularly in sub-Saharan African (SSA) countries, despite increased efforts to control it. The utilization of long-lasting insecticide-treated nets (LLINs) during pregnancy is a well-established strategy to reduce the prevalence of malaria. Nonetheless, inadequate adherence remains a persistent challenge in certain regions with high malaria endemicity. This research aimed to assess the effectiveness of long-lasting insecticidal nets in preventing asymptomatic malaria infections among pregnant women attending antenatal care at the Bonassama District Hospital in the Littoral Region of Cameroon. A hospital-based cross-sectional study was conducted from March to June 2022. Data on sociodemographic characteristics and LLIN usage were collected through a structured questionnaire, while asymptomatic malaria infections were identified using a PfHRP2/pLDH malaria qualitative rapid diagnostic kit. The relationship between categorical variables was analyzed using the chi-square test and logistic regression at a significance level of 5%. Out of the 411 pregnant women included in the study, 156 (35.4%) were diagnosed with malaria. The LLIN utilization rate was 65.1% (287). The risk of malaria infection was 2.7 times higher (AOR = 2.75, 95% CI = 1.83-4.14, p < 0.001) among women who did not consistently use LLINs compared to those who did. The majority (65.1%) of pregnant women always used LLINs, while 34.9% did not consistently use them. Pregnant women aged 20-29 were 3.5 times more likely to use LLINs (AOR = 3.53, 95% CI = 1.61-7.75, p = 0.002) compared to those in the 30-39 and >39 age groups. Furthermore, women practicing Christianity were 1.7 times more likely (AOR = 1.72, 95% CI = 1.01-2.96, p = 0.047) to consistently use LLINs than those of the Islamic faith. Pregnant women in their first trimester (AOR = 3.40, 95% CI = 1.24-4.64, p = 0.010) and second trimester (AOR = 1.90, 95%CI = 0.99 -3.62, p = 0.055) were more likely to sleep under net when compared to those in the third trimester. Among the reasons reported by participants for not frequently using LLINs were heat [55.2% (85)], suffocation [13.6% (21)] and the smell of nets [8.4% (13)]. The use of LLIN was moderately high among the participants in this study, though still below national target. Age group, occupation, religion and gestation period were the major factors determining the use of LLINs. Considering the proven effectiveness of LLINs in reducing malaria morbidity and mortality, it is imperative for the National Malaria Control Programme (NMCP) to remain focused in promoting both LLIN ownership and utilization to achieve the national target of 100% and 80%, respectively.

## Introduction

Malaria due to *Plasmodium falciparum* remains a significant public health challenge, particularly in endemic regions where vulnerable populations such as pregnant women are at increased risk of infection [1]. According to the World Health Organization (WHO), over 249 million cases and 608,000 deaths were recorded in 2022 across the world [2]. Malaria is the most widespread endemic disease in Cameroon. The country is among the 15 highest burden malaria countries, with 2.7% of all global malaria cases and deaths, and 2.3% of malaria deaths occurring in 2021 [3]. The disease burden is disproportionately higher in pregnant women and their newborns who are at increasing risk of severe illness and deaths, with 65 percent of confirmed cases in Cameroon in 2021 being children under five years of age [4,5]. According to the 2022 World Malaria Report, more than 13 million cases of malaria occur in pregnancy globally [2], with Cameroon recording a prevalence rate of 39.8% in 2019 [6]. This in part is due to changes in their immune system, making them more prone to severe complications, including maternal anemia, low birth weight, and maternal mortality [7,8]. A pregnant woman suffering from malaria is estimated to be three times more at risk of dying of the disease compared to a non-pregnant woman suffering from the disease [9]. To mitigate these risks, the use of long-lasting insecticidal nets (LLINs) among other preventive methods, is recommended as a cost-effective and proven intervention to prevent malaria transmission.

Efforts so far put in by the Cameroonian Ministry of health to minimize malaria transmission in pregnancy include the Health Sector Strategy 2016-2027. This has prioritized the fight against malaria with focus on reducing the high maternal and infant mortality through effective control of malaria in pregnancy [3]. The national policy to contain malaria transmission is aligned with the current WHO three-pronged strategic approach in areas of stable *Plasmodium falciparum* transmission which includes Intermittent Preventive Treatment in pregnancy (IPTp), use of Long-Lasting Insecticidal Net (LLIN) and prompt case management [10]. It is worth noting that preventing malaria in pregnancy is also a priority for the Roll Back Malaria (RBM) partnership program [11]. LLINs serve as a physical barrier preventing the mosquito from gaining contact with the body and also kills the mosquito, offering a definitive shield against malaria [12]. Studies carried out in various malaria endemic countries have shown that the effective use of LLINs by pregnant women can reduce the frequency of malaria by half [13–16]. While these endemic countries are benefiting from LLINs use, some do not adhere to this recommendation [17], yet others are still far behind with a low rate of LLIN utilization with consequently, a higher prevalence in pregnant women [18,19]. The probability of LLINs used to reduce the reproduction number *R* (mean population where all individuals are susceptible to infection) of malarial parasites implies that malaria could be eliminated from a community if three quarters of the population uses LLINs[20]. If therefore used properly in endemic areas, LLINs can reduce malaria vector transmission and burden of malaria in the community [12,21].

Despite the proven efficacy of LLINs in preventing malaria, there are persistent challenges related to their utilization among pregnant women. The effective use of LLINS faces several challenges in Cameroon and statistics indicate that they are used only by half of the pregnant women in the country [6]. Despite the mass distribution of LLINs to pregnant women in Cameroon, it is important to note that mere availability may not necessarily lead to their effective utilization as there exist some perceptions and misconceptions about the commodity within the population. Factors such as access to nets, knowledge about their importance, cultural beliefs, and socioeconomic factors can influence the uptake and consistent use of LLINs among this vulnerable population[22,23]. Evaluating the effective use of LLINs, and knowledge that addresses the constraints and promotes positive behaviour is key in preventing malaria in the community [13,24].

Considering the proven effectiveness of LLINs in reducing malaria morbidity and mortality, it is imperative for the National Malaria Control Programme (NMCP) to remain focused in promoting both LLIN ownership and utilization [3,19]. Therefore, understanding the prevalence of malaria among pregnant women and their utilization of LLINs is crucial for targeted interventions to improve maternal and child health outcomes in malaria endemic regions [25,26]. By elucidating the factors associated with both malaria prevalence and LLIN utilization, policymakers and healthcare providers can develop tailored strategies to enhance coverage, promote compliance, and ultimately reduce the burden of malaria in pregnant women [27]. This study aims to investigate the relationship between malaria prevalence and the utilization of LLINs among women attending antenatal clinic at the Bonassama District Hospital. By examining the determinants of LLIN utilization and their impact on malaria infection rates, we seek to inform evidence-based strategies that can enhance the effectiveness of malaria prevention initiatives and ultimately reduce the burden of malaria among pregnant women in Cameroon.

## Materials and Methods

### Study setting and population

This was a hospital based cross-sectional study conducted at the Bonassama District Hospital (BDH) from March 1 to June 30, 2022. The BDH is one of seven health district hospitals in the cosmopolitan city of Douala [28]. The hospital is located in Bonassama Health District which is one of the thirty health districts in the Littoral Region of Cameroon. The district has a surface area of about 55km^2^ with a population of over 350,000 inhabitants and is situated just before the Bonaberi bridge. It attracts patients of diverse socio-economic status living in the city of Douala because of the high quality of services they offer. BDH offers antenatal care to pregnant women, with the hospital being managed by the Director and assisted by the General Supervisor, who coordinates the activities of the hospital.

The study population comprises of women attending antenatal clinic in the health facility during the study period. Any pregnant woman who resided for six months or more prior to data collection period and attending antenatal care at BDH were included in the study. Those who were sick or mentally ill were excluded from the study.

### Sample size and sampling method

The minimum sample size was calculated using the Lorentz’s formula at 95% confidence interval (z), 5% margin of error (d), design effect of 1.5, power of 80% as follows:

N = z^2^pq/d^2^,

Where z^2^ = (1.96)^2^, p = previous malaria prevalence and q = 1-p, d^2^ = (0.05)^2^. PNLP[16] reported a national malaria in pregnancy prevalence (p) of 39.8% in 2019. To compensate for the non-response rate, 10% of the determined sample size was added. The minimum sample size was therefore estimated at 406 women attending ANC at Bonassama District Hospital.

### Data collection

#### Malaria diagnosis

Asymptomatic malaria infection was determined using Bioline rapid diagnostic test (RDT). Capillary blood samples were collected from each participant following a finger prick under standard aseptic procedure. Plasmodium infection status was ascertained using PfHRP2/pLDH malaria rapid diagnostic kit (SD BiolineTM, Alere, South Korea) and results interpreted following manufacturer’s instructions. Briefly, about 5 μl of blood sample from each participant was placed in the sample window of the RDT cassette and three drops of diluent added. The results were then read after 15 minutes, with the presence of two (or three), one or no distinct line indicative of a positive, negative or invalid result, respectively.

#### Survey development

A pre-tested structured interviewer administered questionnaire was used to document information on sociodemographic characteristics of respondents, use of LLINs and source of LLINs. The questionnaire consisted of 11-items. Ten items queried demographic information that included age group, education level, marital status, religion, area of residence, number of children, and gestation period. Four items assessed ownership and utilization of mosquito nets including, source of LLINS, period when the LLINS was gotten, how often the net was used per week, and reasons for not sleeping under the net.

#### Variables

##### Dependent variables

The outcome variables for this study were the usage of LLIN and malaria status. The variable “usage of LLIN” was reduced to binary classes. Sleeping under the net everyday was coded as “Always use LLIN” while “once in a while” or “seldom use” were coded as “Did not always use LLIN”. Malaria status, defined as the result of the asymptomatic malaria qualitative rapid diagnostic test, which was a binary outcome variable was coded as “positive” or “negative”.

##### Explanatory variables

We included possible determinants of LLIN usage previously used in existing literature [17,24,29,30]. The following demographic variables: age group (less than 20, 20-29, 30-39 and above 39), level of education (primary, secondary or tertiary), marital status (single, married or divorced), Religion (Christianity or Islam), place of residence (Bojongo, Bonassama, Mabanda, Sodiko), gestation period (first trimester, second trimester, or third trimester), and gravida (primigravida or multigravida). LLIN related variables such as source of LLIN (purchased or free distribution), period owned LLIN (before ANC or during ANC) were also included.

### Data analysis

All data were entered into Excel and analyzed using SPSS Statistics 26(SPSS Inc, Chicago, USA). Pearson’s χ^2^ test was used to explore significant difference between malaria prevalence and the use of LLIN among pregnant women. Multiple Linear Regression was used to predict malaria prevalence and use of LLINS among pregnant women after controlling for demographic variables. To identify the predictors of use of LLIN, variables with p value of 0.1 or lower in bivariate analysis were included in the logistic regression model at 5% level of significance. Adjusted Odds Ratio (AOR) with 95% confidence interval (CI) was used as a measure of association.

### Ethical consideration

The University of Buea Faculty of Health Sciences Institutional Review Board (IRB), the Littoral Regional Delegation for Public Health and the Bonassama District Hospital approved the study protocol. A written informed consent was obtained from all respondents by way of signing or thumb printing on the informed consent form, after the nature and objectives of this study were explained to them. Participation was completely voluntary. All information collected for the study was treated as confidential and stored in a computer with password protection.

## Results

A total of 441 pregnant women who owned LLIN were enrolled into this study. Most of the women were married (61.0%), within the 20-29 years age group (59.4%), had secondary level of education (51.5%), of Christianity religion (85.7%), and resident at Bojongo (44.0%) (Table 1). The mean age (± SD) of the participants was 27.28 ± 6.95 years. Majority of pregnant women were enrolled in their second trimester (46.5%) of pregnancy and multigravida (81.6%) (Table 1). Most of the pregnant women reportedly had nets before pregnancy (57.6%) and under free distribution (70.1%) either during ANC or by a community health worker (CHW).

**Table 1.** Sociodemographic characteristics, LLIN related factors and malaria prevalence of the study participants (N=441).

| Variable | Category | All participants<br>[n (%)] | RDT Positive<br>[n (%)] | p-value |
| --- | --- | --- | --- | --- |
| Age group | <20 | 29(6.6) | 11(37.9) | 0.416 |
|  | 20-29 | 262(59.4) | 94(35.9) |  |
|  | 30-39 | 108(24.5) | 41(38.0) |  |
|  | >39 | 42(9.5) | 10(23.0) |  |
| Level of education | Primary | 50(11.3) | 13(26.0) | 0.278 |
|  | Secondary | 227(51.5) | 86(37.9) |  |
|  | Tertiary | 164(37.2) | 57(34.8) |  |
| Marital status | Single | 156(35.4) | 49(31.4) | 0.403 |
|  | Married | 269(61.0) | 100(37.2) |  |
|  | Divorced | 16(3.6) | 7(43.8) |  |
| Religion | Christianity | 378(85.7) | 129(34.1) | 0.198 |
|  | Islam | 63(14.3) | 27(42.9) |  |
| Residence | Bojongo | 194(44.0) | 74(38.1) | 0.125 |
|  | Bonassama | 84(19.0) | 34(40.5) |  |
|  | Mabanda | 57(12.9) | 13(22.8) |  |
|  | Sodiko | 106(24.0) | 35(33.0) |  |
| Gestation period | First trimester | 191(43.3) | 70(36.6) | 0.064 |
|  | Second trimester | 205(46.5) | 72(35.1) |  |
|  | Third trimester | 45(10.2) | 14(31.1) |  |
| Gravida | Primigravida | 81(18.4) | 35(43.2) | 0.055 |
|  | Multigravida | 360(81.6) | 121(33.6) |  |
| Source of LLIN | Purchased | 132(29.9) | 46(34.8) | 0.914 |
|  | Free distribution* | 309(70.1) | 110(35.6) |  |
| Period own LLIN | Before pregnancy | 254(57.6) | 82(32.3) | 0.114 |
|  | During ANC | 187(42.4) | 74(39.6) |  |
| Usage of LLIN | everyday | 278 (65.1) | 78(27.2) | <0.001 |
|  | once in a while | 132(29.9) | 59(44.7) |  |
|  | Seldom use | 22(5.0) | 19(86.4) |  |

Free distribution\* = During ANC or by Community health worker (CHW)

### Asymptomatic malaria infection

The prevalence of asymptomatic malaria infection was 35.4% (156/441), with a higher prevalence registered among women within the 30-39 years age group (38.0%, 41/108). The prevalence of malaria infection was generally higher among pregnant women with secondary level of education (37.9%) than their primary (26%) or tertiary (34.8%) level counterpart, though not statistically significant (p> 0.05). Although there was no association between the prevalence of malaria infection and marital status, area of residence, gestation period and gravida, it was generally higher among pregnant women divorced (43.8%), those with Islam religion (42.9%), those residing at Bonassama neighborhood (40.5%) (Table 1). Higher rates of *P. falciparum* infections were also recorded among pregnant women in the first trimester (36.6%), primigravida (43.2%) and among those who received LLIN during ANC (39.4%) (Table 1). Though not statistically significant (p=0.914), rates of asymptomatic malaria infection tended to be higher among pregnant women who received LLIN freely (35.6%) compared to those that purchased LLIN (34.8%). Moreover, malaria prevalence was significantly associated with use of mosquito nets (p<0.001). The infection was higher in pregnant women who rarely slept under the mosquito net (86.4%) compared with those that slept under the net daily (27.2%).

### LLIN usage during pregnancy

Of the 441 LLIN owners, 65.1% of women reported always sleeping under the net, while 29.9% sleep under the net only once in the while. Another 5% seldomly sleep under mosquito nets (Table 1). The primary reason for not always sleeping under the net was due to heat (55.2%). Among other reasons indicated for not always sleeping under the net were, feeling of suffocation (13.6%), windows already have net (9.7%), chemical smell (8.4%), (Fig 1).

**Fig 1.**
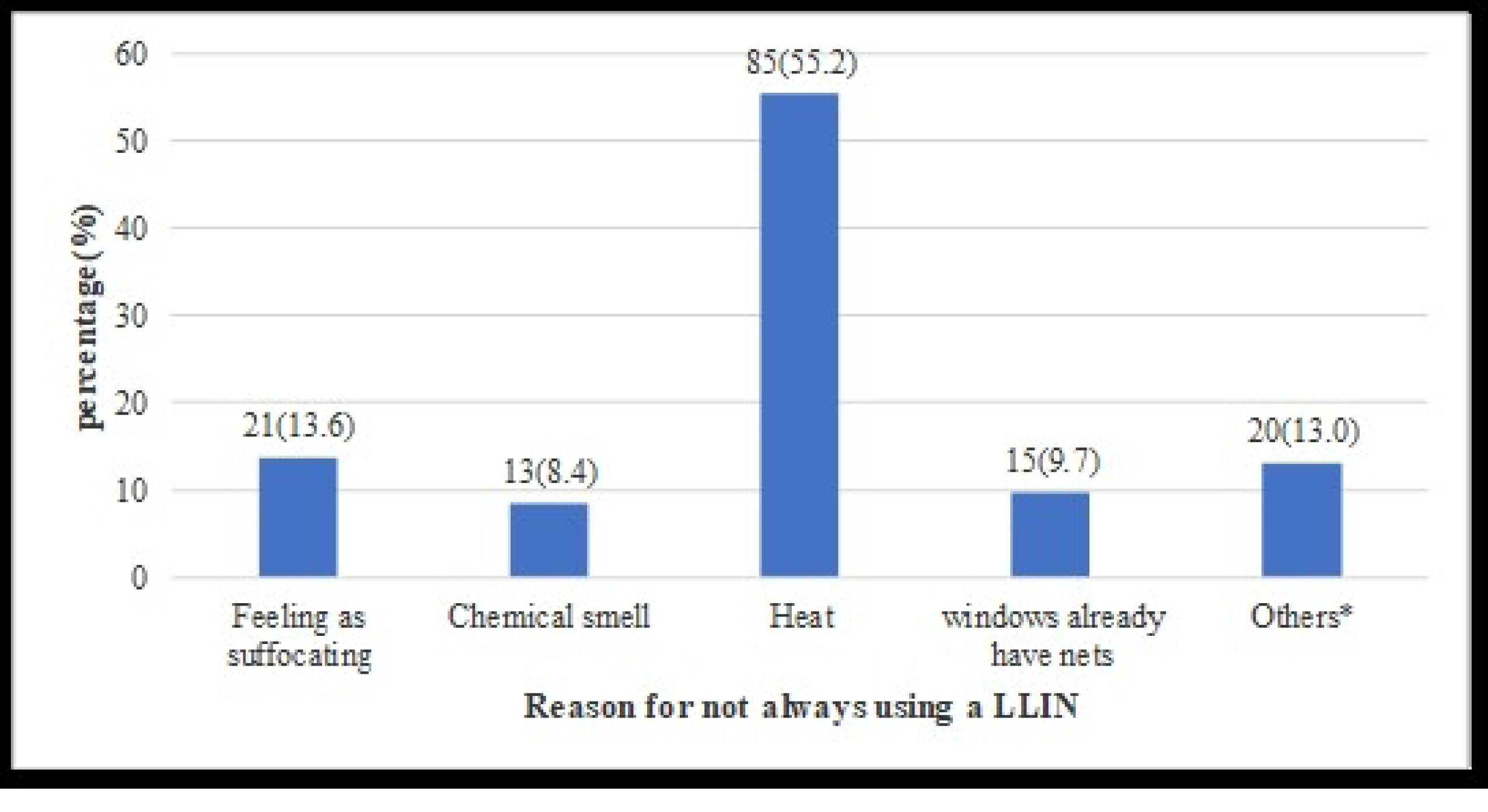
Bed net usage information among women who did not use a LLINS (N=154) *Other Reasons: Allergic reaction (n = 8); Torn net (n = 4); Dirty net (n=6); Irregular mosquitos (n = 2)

To determine the association between sociodemographic factors and use of LLIN, the variable “usage of LLIN” was reduced to binary classes. Sleeping under the net everyday was considered as “Always use LLIN” while “once in a while” or “seldom use” were considered as “Did not always use LLIN”. The majority, 287 (65.1%) of pregnant women always use LLIN, while 154 (34.9%) respondents did not always use LLIN. LLIN usage was independent of level of education, marital status, area of residence, number of children in the family and period in which the net was owned. Nevertheless, LLIN usage was significantly associated with age group, religion, gestation period, source of LLIN and prevalence of malaria infection(p<0.05) (Table 2). Pregnant women in the 20-29 years age category were 3.5 times more likely to use LLIN (adjusted OR = 3.53, 95%CI = 1.61-7.75, p = 0.002) compared to 30-39 and >39 age groups (Table 3). Women of Christianity religion were 1.7 times (adjusted OR = 1.72, 95% CI = 1.01-2.96, p = 0.047) more likely to always use a net compared to Islam religion. However, LLIN usage was significantly associated to gestational age (p=0.031). Pregnant women in their first trimester (adjusted OR = 3.40, 95%CI = 1.24 -4.64, p = 0.010) and second trimester (adjusted OR = 1.90, 95%CI = 0.99 -3.62, p = 0.055) were more likely to sleep under net when compared to those in the third trimester. It was observed that women who did not sleep under LLIN were more susceptible to malaria parasite infection (adjusted OR =2.75-, 95%CI = 1.83–4.14, p < 0.001).

**Table 2.** Relationship between sociodemographic characteristics and LLIN usage.

| Variable | Category | Did not always use LLIN (n=154) | Always use LLIN (n=287) | p-value |
| --- | --- | --- | --- | --- |
| Age group | <20 | 17(58.6) | 12(41.4) | 0.02 |
|  | 20-29 | 75(28.6) | 187(71.4) |  |
|  | 30-39 | 45(41.7) | 63(58.3) |  |
|  | >39 | 17(40.5) | 25(59.5) |  |
| Level of education | Primary | 20(40.0) | 30(60) | 0.67 |
|  | Secondary | 76(33.5) | 151(66.5) |  |
|  | Tertiary | 58(35.4) | 106(64.6) |  |
| Marital status | Single | 56(35.9) | 100(64.1) | 0.68 |
|  | Married | 91(33.8) | 178(66.2) |  |
|  | Divorced | 7(43.8) | 9(56.3) |  |
| Religion | Christianity | 125(33.1) | 253(66.9) | 0.04 |
|  | Islam | 29(46.0) | 34(54.0) |  |
| Residence | Bojongo | 73(37.6) | 121(62.4) | 0.27 |
|  | Bonassama | 22(26.2) | 62(73.8) |  |
|  | Mabanda | 19(33.3) | 38(66.7) |  |
|  | Sodiko | 40(37.7) | 66(62.3) |  |
| Gestation period | First trimester | 58(30.4) | 133(69.6) |  |
|  | Second trimester | 73(35.6) | 132(64.4) |  |
|  | Third trimester | 23(51.1) | 22(48.9) | 0.03 |
| Gravida | Primigravida | 30(37.0) | 51(63.0) |  |
|  | Multigravida | 124(34.4) | 236 (65.6) | 0.693 |
| Source of LLIN | Purchased | 35(26.5) | 97(73.5) |  |
|  | Free distribution | 119(38.5) | 190(61.5) | 0.016 |
| Period own LLIN | Before pregnancy | 29(31.1) | 175(68.9) |  |
|  | During ANC | 75(40.1) | 112(59.9) | 0.05 |

**Table 3:** Logistic regression model for LLIN use among women attending antenatal clinic at Bonassama District Hospital.

| Variable | Category | Always use LLIN (n=287) | Unadjusted p-value | OR | 95%CI | Adjusted p-value |
| --- | --- | --- | --- | --- | --- | --- |
| Age group | <20 | 12(41.4) | **0.003\*** | 1 |  |  |
|  | 20-29 | 187(71.4) |  | 3.53 | 1.61-7.75 | **0.002\*** |
|  | 30-39 | 63(58.3) |  | 2.00 | 0.86-4.56 | 0.107 |
|  | >39 | 25(59.5) |  | 2.08 | 0.80-5.45 | 0.135 |
| Level of education | Primary | 30(60) | 0.675 | 1 |  |  |
|  | Secondary | 151(66.5) |  | 1.32 | 0.71-2.49 | 0.381 |
|  | Tertiary | 106(64.6) |  | 1.22 | 0.64-2.33 | 0.551 |
| Marital status | Single | 100(64.1) | 0.685 | 1.39 | 0.49-3.93 | 0.536 |
|  | Married | 178(66.2) |  | 1.52 | 0.55-4.22 | 0.420 |
|  | Divorced | 9(56.3) |  | 1 |  |  |
| Religion | Christianity | 253(66.9) | **0.049\*** | 1.72 | 1.01-2.96 | **0.047\*** |
|  | Islam | 34(54.0) |  | 1 |  |  |
| Residence | Bojongo | 121(62.4) | 0.275 | 1.00 | 0.62-1.64 | 0.985 |
|  | Bonassama | 62(73.8) |  | 1.71 | 0.91-3.19 | 0.093 |
|  | Mabanda | 38(66.7) |  | 1.21 | 0.62-2.34 | 0.577 |
|  | Sodiko | 66(62.3) |  | 1 |  |  |
| Gestation period | First trimester | 133(69.6) | **0.031** | 3.40 | 1.24-4.64 | **0.010\*** |
|  | Second trimester | 132(64.4) |  | 1.90 | 0.99-3.62 | 0.055 |
|  | Third trimester | 22(48.9) |  | 1 |  |  |
| Gravida | Primigravida | 51(63.0) | 0.693 | 1 |  |  |
|  | Multigravida | 106(69.3) |  | 1.12 | 0.68-1.85 | 0.658 |
| Source of LLIN | Purchased | 97(73.5) | **0.016\*** | 1.74 | 1.11-2.72 | **0.016\*** |
|  | Free distribution | 190(61.5) |  | 1 |  |  |
| Period own LLIN | Before pregnancy | 175(68.9) | 0.050 | 1.48 | 1.00-2.20 | 0.050 |
|  | During ANC | 112(59.9) |  | 1 |  |  |

**OR = Odds Ratio; CI = Confidence Interval; 1= Reference group; \*Significant p values**

### LLIN usage and risk of malaria infection

Of the 441 pregnant women who owned LLIN, the rate of malaria infection was 27.7% (78/287) and 50.6% (78/154) for those who always use and do not always use LLIN, respectively. LLIN usage was significantly associated with malaria infection (p<0.001). However, the risk of malaria infection was 2.7 times (adjusted OR = 2.75, 95%CI = 1.83-4.14, p <0.001) higher among those who did use LLIN compared to those who always used LLIN (Table 4).

**Table 4:** LLIN usage and risk of malaria infection.

| LLIN usage | Malaria prevalence<br>[(%) n/N] | Unadjusted<br>p-value | OR | 95% CI | Adjusted<br>p-value |
| --- | --- | --- | --- | --- | --- |
| Not always use LLIN | (50.6) 78/154 | <0.001 | 2.75 | 1.83-4.14 | <0.001 |
| Always use LLIN | (27.2)78/287 |  | 1 |  |  |

## Discussion

Though LLINS have been shown to reduce maternal and child malaria related morbidity and mortality, their effective use remains a problem in malaria endemic regions. Measures to promote their effective utilization are crucial in improving maternal and child health outcomes. The current study was conducted to evaluate the prevalence of malaria infection and the use of mosquito bed net among pregnant women in Bonassama District Hospital in the Littoral Region of Cameroon. The prevalence of asymptomatic malaria infection among pregnant women in this study was 35.4% (156/441). The result corroborates other studies conducted in malaria endemic regions in Nigeria (41.6%) [9] and Ghana (42%) [31]. However, the observed prevalence was lower when compared to the 39.8% national prevalence obtained in Cameroon in 2019[6], and other studies conducted in Cameroon [32] and Nigeria [33] reporting an asymptomatic malaria prevalence of 82.4% and Nigeria 79.5%, respectively. On the other hand, the finding of this study was much higher than primary studies in Bangladesh (2.3%) [34], Cameroon (10.1%) [16], a meta-analysis in Nigeria (23.4%) [33] and primary study in Burkina Faso (23.9%) [35]. The Littoral Region of Cameroon is characterized by poor drainage/sewage disposal systems and flooding which are suitable breeding grounds for malaria vectors which contribute to the high prevalence observed in this study. Moreover, the high prevalence reported in this study may also be due to multiple factors including seasonal changes, intensity of the transmission, adherence to malaria preventive measures and the type of diagnostic test done. Overall, this result emphasis the need to include routine laboratory diagnosis of asymptomatic malaria infection as part of antenatal care follow-up for early detection and treatment to prevent negative effects due to malaria infection.

In this study, pregnant women who were in the first trimester of pregnancy were at risk of developing malaria infection compared to women in the second and third trimester. Moreover, women who were primigravida have increased risk of malaria infection compared to multigravida. Similar results were found from studies conducted in Gabon [36], Ghana [21], Ethiopia [37] and Cameroon [29]. Miller et al [38] suggested that the high prevalence of malaria observed during the first trimester may due to the adherent proteins on the surface of infected red blood cells (IRBCs), enabling the IRBCs to adhere to microvascular capillaries of vital organs causing severe parasitological condition. In addition, it may also be due to the fact that pregnant women usually do not attend antenatal visits early in pregnancy and a large proportion of them might have unrecognized and untreated malaria infection as most infections are asymptomatic [39]. The reason for primigravida having a higher risk and burden of asymptomatic malaria infection than multigravida could be related to parity level immunity to variant surface antigen of P. falciparum erythrocyte membrane protein 1 (PfEMP1), which is acquired through consecutive pregnancies [40].

The result from this study indicates that majority of pregnant women (65.1%) used LLIN as a measure to prevent malaria in the current pregnancy. This result is consistent with research from Zambia (68%) [41], South Eastern Nigeria (70%) [42], Uganda (73%) [17] with some studies from Ghana (94.8%) [43] and Rwanda (87.6%) [30] having higher rates of utilization. The varied socioeconomic situations, geographic locations, and approaches to malaria control in the aforementioned nations may be the reason for these differences. These percentages are however higher than the 58.3% [24], 57.8% [16] and 42.7% [29] reported in the South West, North West and West regions of Cameroon, respectively. Though the utilization of LLIN by participants of this study was relatively high, it still falls below the national target of 80%, suggesting the need for continuous sensitization on effective utilization of LLINS.

Furthermore, this study identified factors associated with the use of LLINs among pregnant women. Age group was found to be significantly associated with the use of LLINs. The odds of using LLINs for pregnant women who were older (>39 years) were 2 times higher than those who were younger (<20 years). This is supported by research conducted in northern Ethiopia [44] and Rwanda [30] which reported that old-aged women were more likely to utilize LLINs compared to their counterparts. Moreover, in our study, pregnant women who were Christians were 2 times more likely to sleep under LLIN than their Muslim counterpart. The findings of religion as a possible determinant of LLIN use is supported by studies conducted in Ghana [45,46]. Dun-Dery et al [45] suggested that this could be due to the fact that Muslim men are less supportive to their spouses. The study also revealed that pregnant women who were in their first trimester were 3 times more likely to use their LLINs than those in the third trimester. who registered for ANC during their first trimester. In this study pregnant women who purchased LLINs were more likely to use LLIN than those who had received them freely. Free delivery of LLINs has been shown not to necessarily increase LLIN use [23]. Some studies have shown that personal decision to purchase a net may motivate one to use it rather than getting it for free [47]. This study found that the level of education, marital status, area of residence, gravida, and period in which the participants own the LLIN were not associated with use of LLINs.

In this study, not using LLIN increases the odds of developing malaria infection during pregnancy. In fact, the prevalence of asymptomatic malaria infection was 3 times higher in those that do not always used LLIN than those who always used LLIN. This study’s finding was in agreement with the study conducted in the South West Region of Cameroon [24], Ethiopia [19,37], Malawi and Nigeria [18] which showed that the use of bed nets has a significant impact on decreasing malaria infection. This therefore shows that the use of LLINs as community-level intervention does not only directly prevents the mosquito from biting an individual, but has a lethal effect on the mosquito as well [48], thus reduces the mosquito infestation at household and community levels since more protection is gained from one or more ITNs in each household [49].

## Limitations

Our study was a cross-sectional study design, it does not show a direct temporal relationship. Moreover, though using PCR and microscopy may have higher sensitivity in the diagnosis of malaria infection, we could not do these tests therefore, the result of this study could be affected by the inherent performance of the RDT utilized. In addition, with all survey data, the findings are limited by recall and social desirability biases. Some answers to questions such as sleeping under LLINs, were reported by the participants not observed by the researcher. Likewise, it was not possible for the team to verify the status of nets during the survey. Finally, this study cannot be generalized to the whole population in Cameroon as the sample was selected in only one health facility.

## Conclusion

In this study, the prevalence of asymptomatic malaria among pregnant women was found to be higher than the national average, albeit lower than expected. Factors such as age group, religion, duration of ownership, and consistent use of LLINs showed significant associations with malaria infection. The utilization rate of LLINs was moderately high and was notably linked to age group, religion, gestational period, and the source of LLINs. These findings underscore the importance of conducting ongoing awareness campaigns to promote both ownership and proper usage of LLINs. Additionally, there is a clear need to incorporate routine laboratory screening for asymptomatic malaria into antenatal care protocols to enable early detection and treatment, thus minimizing the adverse impacts of malaria infection.

## Data Availability

The authors will made available without any restrictions all data underlying the findings.

## Acknowledgments

We would like to thank the study participants who made this study realistic, and the Director of the Bonassama District Hospital for the permission granted to conduct the study in the health facility.

## Author Contributions

**Conceived and designed the experiments:** Marcelus U. Ajonina, Tobias O. Apinjoh

**Data curation:** Marcelus U. Ajonina, Carine K. Nfor, Derick N. Awambeng

**Formal Analysis:** Marcelus U. Ajonina, Tobias O. Apinjoh, Irene U. Ajonina-Ekoti

**Contributed reagents/materials/analysis tools:** Marcelus U. Ajonina, John Ngulefac

**Wrote the paper:** Marcelus U. Ajonina, Irene U. Ajonina-Ekoti, John Ngulefac, Martin Ayim

